# Two-Stage Bayesian Factor Analysis for Air Pollution Source Apportionment and Health Risk Assessment

**DOI:** 10.1101/2025.11.05.25339564

**Authors:** Georges Bucyibaruta, Monica Pirani, Christina Mitsakou, David Green, Gary W. Fuller, Anja Tremper, Marta Blangiardo

## Abstract

Air pollution, especially particulate matter (PM), presents significant public health challenges and is associated with several Sustainable Development Goals (SDGs), notably SDG 3 (Good Health and Well-being) and SDG 11 (Sustainable Cities and Communities). Effective policy development requires robust statistical approaches to identify pollution sources and quantify their health impacts with appropriate uncertainty.

This study develops a novel two-stage Bayesian framework for air pollution source apportionment and health risk assessment, with the aim of quantifying the contribution of distinct particle sources to respiratory health outcomes in children, using particle number size distribution (PNSD) data from London.

In the first stage, we construct a Bayesian dynamic factor model with autoregressive components to infer latent pollution sources, incorporating non-negativity constraints and accounting for temporal dependence. In the second stage, we assess the relationship between source-specific exposures and respiratory hospital admissions in children via a Poisson regression model, explicitly propagating uncertainty from the source apportionment stage to the health model.

The model identifies four main sources: nucleation, traffic, urban activities, and secondary aerosols. Among these, traffic and secondary sources exhibit the strongest and most consistent associations with increased respiratory hospital admissions. Importantly, models that do not account for uncertainty propagation tend to overestimate health risk associations, underscoring the value of the proposed Bayesian framework.

This work illustrates the advantages of integrating Bayesian methods for source apportionment and health effect estimation, with formal uncertainty propagation across model stages. The proposed framework enhances interpretability and supports evidence-based public health and environmental policy. It is readily extensible to other pollutants and settings, contributing to improved air quality management and progress toward global sustainability goals.

## 1 Introduction

Air pollution is a major public health concern, and there is extensive evidence of its role in a range of non-communicable diseases, especially respiratory and cardiovascular diseases (Evangelopoulos et al., 2019; Fuller, 2019; Fuller et al., 2022). Air pollution is addressed in several Sustainable Development Goals (SDGs), including SDG 3 (Good Health and Well-being), SDG 11 (Sustainable Cities and Communities), and SDG 13 (Climate Action), highlighting its impact on health, urban environments, and climate systems (WHO, 2025). In the UK, the Committee on the Medical Effects of Air Pollutants (COMEAP)^1^ has been reviewing the toxicological and epidemiological evidence, advising the government and informing the research needs on the health effects associated with exposure to air pollution.

Particulate matter (PM), defined as any particle smaller than a certain size (e.g. PM_10_, PM_2.5_) has been recognised as one of the most dangerous pollutants (WHO, 2021). PM contains a mixture of contaminants from different sources, nevertheless the majority of epidemiological studies are still considering the totality of PM when assessing a link with health outcomes. Several recent studies have questioned whether a more meaningful approach is to studying the source composition of PM concentration, to account for the fact that different sources might have different harmful effects (see for instance Park et al. 2014; Hackstadt and Peng 2014; Pirani et al. 2015; Samoli et al. 2016; Boogaard et al. 2022; COMEAP 2022). This perspective also allows policy-makers to more directly connect research outcomes to interventions.

In this perspective, source apportionment (SA) refers to a class of methods aimed at partitioning pollution to the sources from which it was emitted. SA has been an active area of research in the last thirty years (Hopke, 2016; Yu et al., 2022). Two main approaches can be considered: i) source-oriented deterministic models; and ii) receptor models (Viana et al., 2008). The former relies on knowledge of emissions and transport processes to predict the concentrations of pollutants at a receptor site; the latter is based on statistical procedures to identify and quantify the sources of pollutants based on the mixture of chemicals measured at the receptor sites. Receptor models have become popular as they do not require prior knowledge on the emission sources; additionally, software for this type of analysis is widely available.

To perform SA, compositional data with information on the different chemical components within PM concentration are needed to disentangle the total PM into sources. However, this type of data is expensive to collect and not widely available at monitoring site networks. As an alternative, measurement of particle number concentration (PNC) and related particle number size distribution (PNSD) have recently received much attention as a way to investigate PM sources (Hopke, 2016). This consists of measuring a wide range of particle sizes, ranging from 10nm to 2500nm diameter, spanning both ultrafine (< 100nm diameter) and fine (100 - 2500nm diameter) particle ranges. Similar results have been reported when comparing sources from compositional data and PNSD, particularly for long-range transport sources (Gu et al., 2011). In the UK, Martínez-Hernández et al. (2025) proposed a functional factor model for hourly PNSD collected at a background station in Greater London, while Baerenbold et al. (2023) proposed a dependent Dirichlet process mixture model to apportion PNSD collected at a monitoring station near London Gatwick airport. Other studies relied on the popular Positive Matrix Factorization (PMF) to separate airborne PNSD into sources (Beddows et al., 2015; Beddows and Harrison, 2019; Tremper et al., 2022). Furthermore, a recent study applied PMF to analyze the air pollution mixture in four European cities, including London, identifying common sources of PNSD in cities while also distinguishing location-specific ones (Rivas et al., 2020).

A recent review on receptor modelling methods for air pollution SA, underlined that deterministic models do not provide accurate representations of the variability of species and concentrations observed in the atmosphere (Hopke, 2016), while other authors have recommended the use of a Bayesian approach to overcome this issue (Park et al., 2015; Molitor et al., 2016; Pirani et al., 2015). For example Hackstadt and Peng (2014) was one of the first studies to propose a Bayesian factor model for source attribution, using national databases to provide prior information on source emissions.

The Bayesian paradigm has also been advocated as a natural framework for ensemble-based SA methods, which try to overcome limitations and uncertainty associated with individual standard SA models (Park and Oh, 2015; Tang et al., 2020), along with specific computational packages (Park et al., 2021). These contributions highlighted the need for additional work to better understand the potential of Bayesian approaches in receptor modelling. For instance, Park et al. (2001) included in the model an autoregressive component to account for temporal correlation in the data, while Park and Oh (2015) explicitly accounted for nonnegativity constraints on the source contributions and source compositions, in both parameter and model uncertainty estimation.

An additional and related challenge arises when evaluating the health effects of pollutant mixtures. The typical epidemiological approach consists of two-stages: the source profiles are estimated, and then they are plugged into a health regression model (see for instance Samoli et al. 2016; Bell et al. 2014; Rivas et al. 2021). Recently, statisticians have been advocating that uncertainty in the exposure estimates needs to be taken into account in the health-effect model. While several contributions have considered a joint approach, where the source profiles are estimated at the same time as their impact on health outcomes (Park et al., 2014; Molitor et al., 2016), this approach has been criticised as it potentially introduces feedback from the health endpoint into the source estimation (Szpiro and Paciorek, 2013).

Framed in this perspective, here we propose a two-stage model to apportion PNSD; the first stage allows for a data driven selection of the number of sources based on prior knowledge of sources characterization, and explicitly accounts for nonnegativity constraints on the source contributions and source compositions. Additionally, it includes an autoregressive component to account for temporal correlation in the data. The second stage links the apportioned sources to health outcomes; in particular, we evaluate the effect of the estimated sources on respiratory hospital admissions in children. To propagate the uncertainty from the first stage (SA) to the second stage (health model), we select a number of samples from the first stage, to represent the sources posterior multivariate distribution; we then run the second stage and combine the effect estimates using weights based on the likelihood of each of the selected first stage samples. We finally compare the results with that of a classic two-stage model which plugs-in point estimates only from the first stage.

The remainder of the paper is structured as follows: in section 2 we present the data and the proposed statistical approach. Then, in section 3, we describe the results of the two-stage model, including the choice of the number of sources and their interpretation, as well as comparing the epidemiological results when we include or do not include the uncertainty propagation. Finally, in section 4 we provide some discussion points on the model specification, on the results and on the policy implications.

## 2 Material and methods

### 2.1 Data

#### 2.1.1 Sampling site and PNSD data

The air quality data analysed in this study consist of PNSD that have been measured during the years 2014-2016. These PNSD data were collected from an urban background monitoring site located in a residential area of London (North Kensington, 51° 31′ 16′′ North, 0° 12′ 48′′ West). This station is part of both the London Air Quality Network and the national Automatic Urban and Rural Network, and is located on the grounds of Sion Manning School. The closest road, St Charles Square, is a quiet residential street located approximately 5 m from the monitoring site, within a predominantly residential area. The nearest major paved roads are the B450, 100 m to the east, and the heavily trafficked A40, 400 m to the south. Data were acquired using a Scanning Mobility Particle Sizer (SMPS; TSI 3080) coupled with a Condensation Particle Counter (CPC; TSI 3775) equipped with a long Differential Mobility Analyzer (DMA) and a diffusion dryer. These measurements adhered to the EUSAAR/ACTRIS protocol recommendations (Wiedensohler et al., 2012) and were subsequently corrected for diffusional losses. PNSD data were measured over 51 size bins, ranging between 17 and 604 nm. For this study, we aggregated the hourly data to obtain mean daily time series.

#### 2.1.2 Gaseous pollutants and meteorological data

We collected daily time series concentration data for various gaseous pollutants, including carbon monoxide (CO; ppb), nitrogen oxides (NO_x_; ppb), nitrogen dioxide (NO_2_; ppb), nitric oxide (NO; ppb), ozone (O_3_; ppb), and sulfur dioxide (SO_2_; ppb), using the R package `openair` (Carslaw and Ropkins, 2012). Meteorological data, such as daily mean temperature (°C), atmospheric pressure (hPa), visibility (m), and relative humidity (%), were also obtained via the same package. Black carbon (BC_IR_; ppb and BC_UV_; ppb) are measured through the magee aethalometer two wavelength at the North Kensington monitoring station, while BC_wood_ (ppb) is computed from the previous two measurements following Font et al. (2022). In addition, wind speed (m/s) and direction (angle) data were retrieved from the NOAA Integrated Surface Database for the London Heathrow site using the R package `worldmet` (Carslaw, 2023).

#### 2.1.3 Hospitalization data

The data related to hospital admission for respiratory diseases (ICD 10 codes: J00–J99) among children (<16 years of age) were available from the registry of the Hospital Episode Statistics within the Small Area Health Statistics Unit (SAHSU) of Imperial College London and cover all children resident within the Greater London boundaries.

### 2.2 Two-stages modelling approach

Our modelling approach consists of two stages. In the first stage, we specify a dynamic factor model to estimate the latent air particles sources and their associated uncertainty. In the second stage we link the identified pollution sources to the number of hospital admissions for respiratory causes in children in Greater London.

The two-stages model is specified in details in the following sections. Both stages are implemented using an MCMC algorithm within the rjags package (Finley, 2013) in R version 4.2.1. For the first stage we considered 2 chains with 50000 iterations as burnin and 50000 as posterior sample thinned by 10, and 10000 as adaptation sample. For the second stage we considered 2 chains with 40000 iterations as burnin and 10000 as posterior distribution, thinned by 20, so that the final posterior sample was 1000 across the two chains. The convergence to the stationary distribution was evaluated using traceplots and the Rhat analytical tool (Gelman et al., 2013).

#### 2.2.1 Bayesian dynamic factor model for SA

We propose a dynamic multivariate receptor model to estimate PNSD source profiles and contributions. Let *X*_*t*_ = (*X*_*t*1_, *X*_*t*2_, …, *X*_*tD*_) be a vector of particle size bins of dimension *D* measured at time *t*, for *t* = 1, …, *T*. The model is specified as:

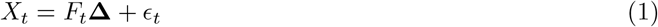

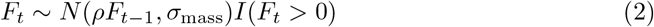

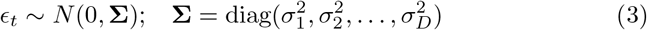

with *F*_1_ ∼ *N* (0, *σ*_*f*_)*I*(*F*_1_ > 0). The interpretation of terms in Eq. (1) is similar to Park et al. (2014), i.e., *F*_*t*_ = (*F*_*t*1_, …, *F*_*tK*_) is the source contribution vector in day *t* (*t* = 1, …, *T*) from each unobserved source type *k* (*k* = 1, …, *K*) while **Δ** represents the source composition matrix of dimension *K* × *D*, where the rows are the source composition profiles, Δ_*k*_ = (Δ_*k*1_, …, Δ_*kD*_), consisting of the relative contribution of each size bin to each source *k*. Lastly, *ϵ*_*t*_ = (*ϵ*_*t*1_, …, *ϵ*_*tD*_) represents the measurement error. To ensure model identifiability, we consider three constraints as specified in Park et al. (2021): (i) there are at least *K*−1 zero elements in each row of **Δ**; (ii) for each *k* = 1, …, *K*, the rank of Δ(*k*) is *K* − 1; (iii) 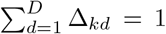 for each *k* = 1, …, *K*. Differently from Park et al. (2014, 2021) in Eq. (2) we allow the unobserved factors *F*_*t*_ to evolve in time according to an autoregressive process of order 1 (AR1) with parameter |*ρ*| < 1, acknowledging the temporal autocorrelation structure intrinsic in the time series.

To complete the Bayesian model, the prior distributions for the unknown parameters need to be specified. Following Park et al. (2021), for **Δ**, we assume a point mass at zero for *K*(*K* − 1) elements, pre-selected for enforcing identifiability condition *C*_1_. For the free elements of **Δ**, we consider the truncated Normal distribution, *vec*Δ^+^ ∼*N*_*DK*−*K*(*K*−1)_(*µ*_0_, Σ_0_)*I*(*vec*Δ^+^ *≥*0). Note that *vec*Δ^+^ denotes the *DK* −*K*(*K* − 1)-dimensional vector of free elements of Δ stacked columnwise, to incorporate the non-negativity constraints, critical to ensure that the estimated source profiles have a reasonable interpretation from a physical-chemical sense, while facilitating computation. The parameter *µ*_0_ is an *DK* − *K*(*K* − 1)-dimensional vector and Σ_0_ is an (*DK* − *K*(*K* − 1) × *DK* − *K*(*K*−1))-dimensional diagonal matrix and their elements are prespecified. Out of the total number of elements in *µ*_0_, some need to be set to zeros to satisfy the identifiability condition C1. We used information from previous studies (e.g. Rivas et al. (2020) to select bins not compatible with the *k* source and putting these values equal to 0. For the remaining elements, any nonnegative numbers (between 0 and 1 preferably, Park et al. (2014)) can be assigned; we fixed those values to 0.5.

Additionally, we assume the common inverse gamma prior for the diagonal elements of 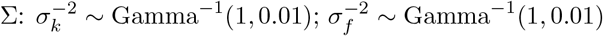 and 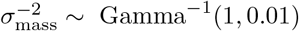, while the autoregressive parameter *ρ* ∼ Uniform(0, 1).

#### 2.2.2 Selection of the number of sources

The number of sources *K* needs to be defined a priori; the common approach consists of running multiple models with different *K* and then inspect the sources in terms of temporal profiles and correlation with other gasses (e.g., O_3_, NO_2_) and meteorological conditions (e.g., temperature, wind speed and direction) to choose the most ‘reasonable’ configuration. Here we employ an alternative principled approach, by means of two criteria: the posterior Bayesian *R* squared and the Bayesian p-value.

We use *X* to denote the observed data, *M*_*g*_ to identify the model, and ***θ*** for the vector of unknown model parameters. For each draw of ***θ***_*i*_, (***θ***_*i*_, *i* = 1, …, *m*), we compute a vector of *L* = *T* × *D* predicted values 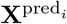 from the posterior predictive distribution. Then the explained variance *V* is calculated as:

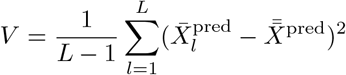

while the residual variance var_res_ is calculated as:

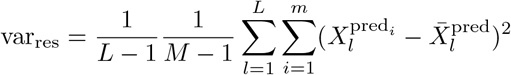

and thus the posterior distribution of the Bayesian *R*^2^ is:

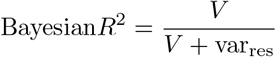

We also calculate a posterior p-value (Gelman et al., 2013), defined as the probability, given the data, that a predicted observation is smaller than the observed data. Given a set of draws, *i* = 1, …, *m*, we calculate the following for each *i*:

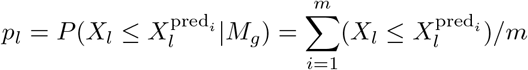

which estimates the proportion of the *m* draws for which 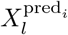 exceeds *X*_*l*_. Having obtained *p*_*l*_, *l* = 1, …, *L*, we visualise it through an histogram, as recommended in (Gelman et al., 2013). If the model represents the data well, the histogram should resemble a uniform distribution, while deviation from it points towards model mispecification.

#### 2.2.3 Health model

In the second stage, we link the estimated sources to health data through a Poisson regression model. This stage assesses the association between sources and hospital admissions for respiratory causes in children through the following specification (which from now on we refer to as the **health model**):

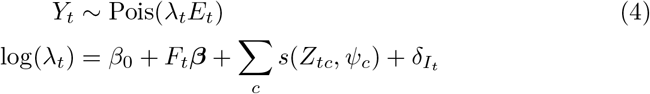

where *Y*_*t*_ is the health outcome at time *t*; *β*_0_ is the general baseline risk of health outcome; ***β*** = (*β*_1_, …, *β*_*K*_) is the vector of parameters describing the effect of each source-specific exposure on the risk of hospitalization for respiratory conditions; *s*(*·, ψ*_*c*_) denote nonlinear functions of daily average temperature and time, which account for seasonality and long-term trend. Finally, the term *I*_*t*_ is an indicator variable that classifies the days of the week according to workday or holiday. We used flexible nonparametric penalised spline functions (Ruppert et al., 2003; Crainiceanu et al., 2005) to capture the nonlinear effect of temperature and time, specifying a low-rank thin plate spline, similarly to Pirani et al. (2016); Blangiardo et al. (2019), as it tends to show a smaller posterior correlation between parameters compared to other types of splines. By letting *Z*_*tc*_ be the *c*-th confounder on day *t*, the spline specification in Eq. (4) can be extended to:

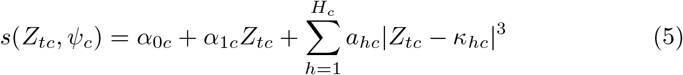

where ***α***_**0**_ and ***α***_**1**_ are fixed unknown parameters, and ***a*** = (*a*_1_, *a*_2_, …, *a*_*H*_)′ is a vector of random parameters corresponding to the set of basis functions *Z*_*tc*_ − *κ*_*hc*_ for the cubic splines. However, given that the *c* separate intercept terms ***α***_**0**_ will not be identified, we dropped those and only took one global intercept as in Eq. (4) (Pirani et al., 2016). The knots of the splines are chosen at equally spaced quantiles; we selected four knots for temperature and nine for time, after preliminary exploratory analysis.

The priors for the health model were specified as follows: *β*_0_ ∼ *N* (0, 1); *β*_*k*_ ∼ *N*_*K*_(0, 1); *δ* ∼ *N* (0, 1) while a hierarchical specification is assigned to the coefficients of the basis functions, that is, 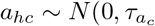 with proper prior on the precision parameter (inverse of variance) 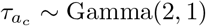, which ensure penalisation.

### 2.3 Uncertainty propagation

An important aspect of the model consists of the propagation of the uncertainty on the apportioned sources (first stage) into the health model (second stage). In this context, we explored two approaches; in the first we estimated source-specific health effects by plugging in the posterior mean of the *F*_*t*_ in the health model (called **plug-in** model). This is the approach typically used in environmental epidemiology, consisting in synthesizing the posterior distribution of the exposure through the posterior mean and then plugging this in the health model (see for instance Rivas et al. (2021)). The second approach takes advantage of the whole posterior distribution of *F*_*t*_ generated in stage one and employs the following steps:

1. starting with a sample from the posterior distribution for the matrix *F*, so that we have *I* vectors with length *T* × *K*, for *i* = 1, …, *I* we calculate the joint posterior density using a Normal approximation 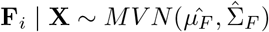 with 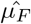 and 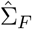 the posterior mean vector and variance-covariance matrix.
2. We synthesize the *I* values into a computationally more manageable number *G* using the quantiles of the densities **w** = (*w*_1_, …, *w*_*i*_, …, *w*_*I*_), with *w*_*i*_ = *f* (**F** = **F**_*i*_ | **X**) to ensure that we consider the whole spread of the distribution.
3. We run the health model *G* times, each one selecting *F* as the matrix of posterior values where the joint density corresponds to *w*_*g*_, *g* = 1, …, *G*.

While step 1-3 are in line with previous work on uncertainty propagation in two-stage models (see for example Blangiardo et al. 2016; Lee et al. 2017; Villejo et al. 2023), here we combine each ***β*** = {*β*_1_, …, *β*_*K*_} using a weighted sum of the *G* runs, each weighted by the *w*_*g*_ value (**weighted approach**).

Additionally, we considered a **unweighted** approach, which consists of combining the posterior distribution for the vector of ***β*** from each of the the *G* samples all with equal weights. This is the approach used in Blangiardo et al. (2016); Lee et al. (2017); Villejo et al. (2023).

## 3 Results

Figure 1 plots the correlation matrix across bin sizes, showing a strong dependence from bins close in size, with values even larger than 0.97 (Figure 1). This stresses the importance of considering bin size dependence in the model formulation.

**Figure 1.**
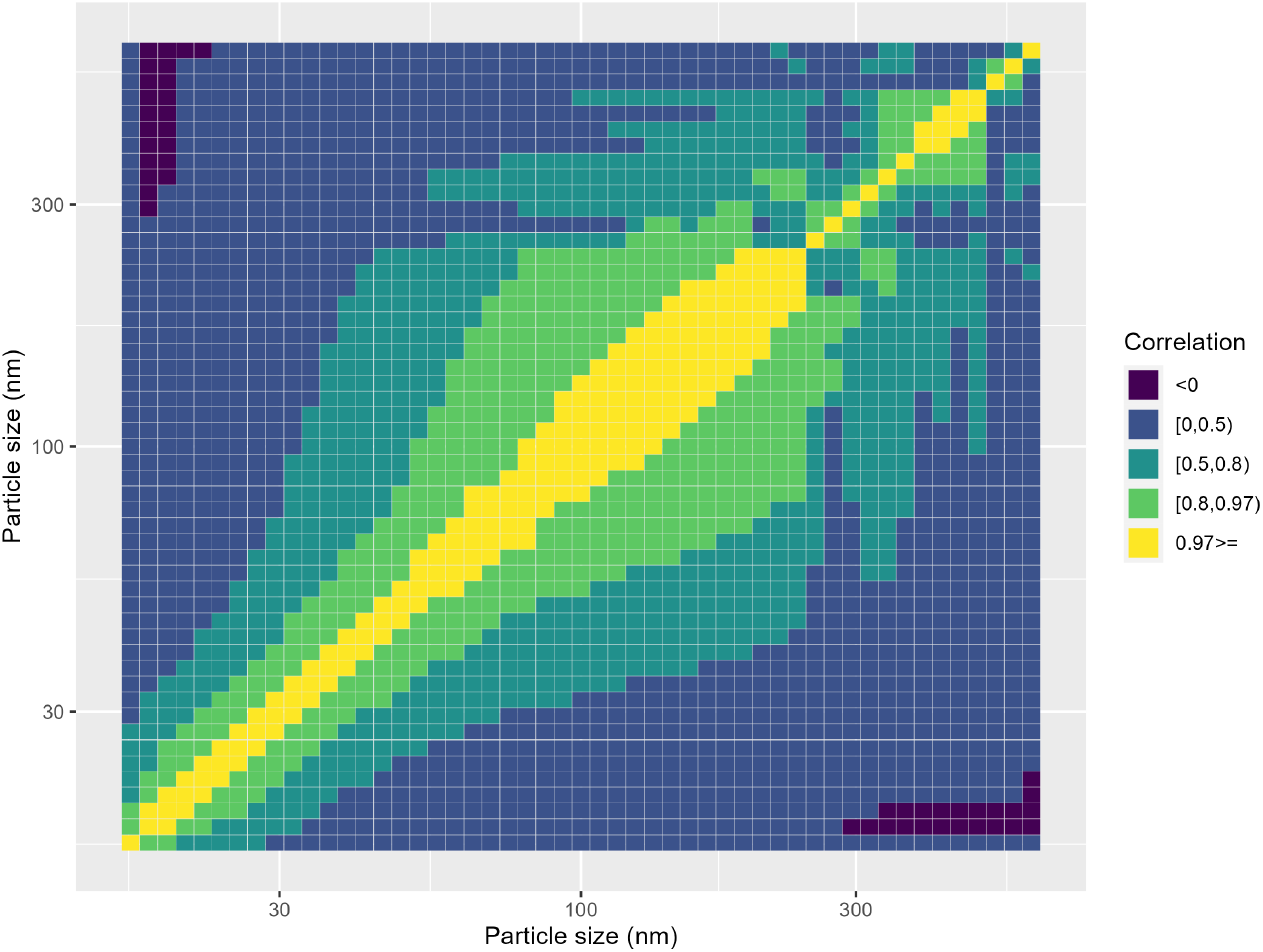
Correlation matrix for bin sizes

The daily paediatric hospitalisations for respiratory causes range from 20 to 221 in the study period, with an average of 92.6 which we used as *E*_*t*_ in the health model. There is a clear seasonal pattern and a nonlinear relationship with temperature as shown in Figure 2.

**Figure 2.**
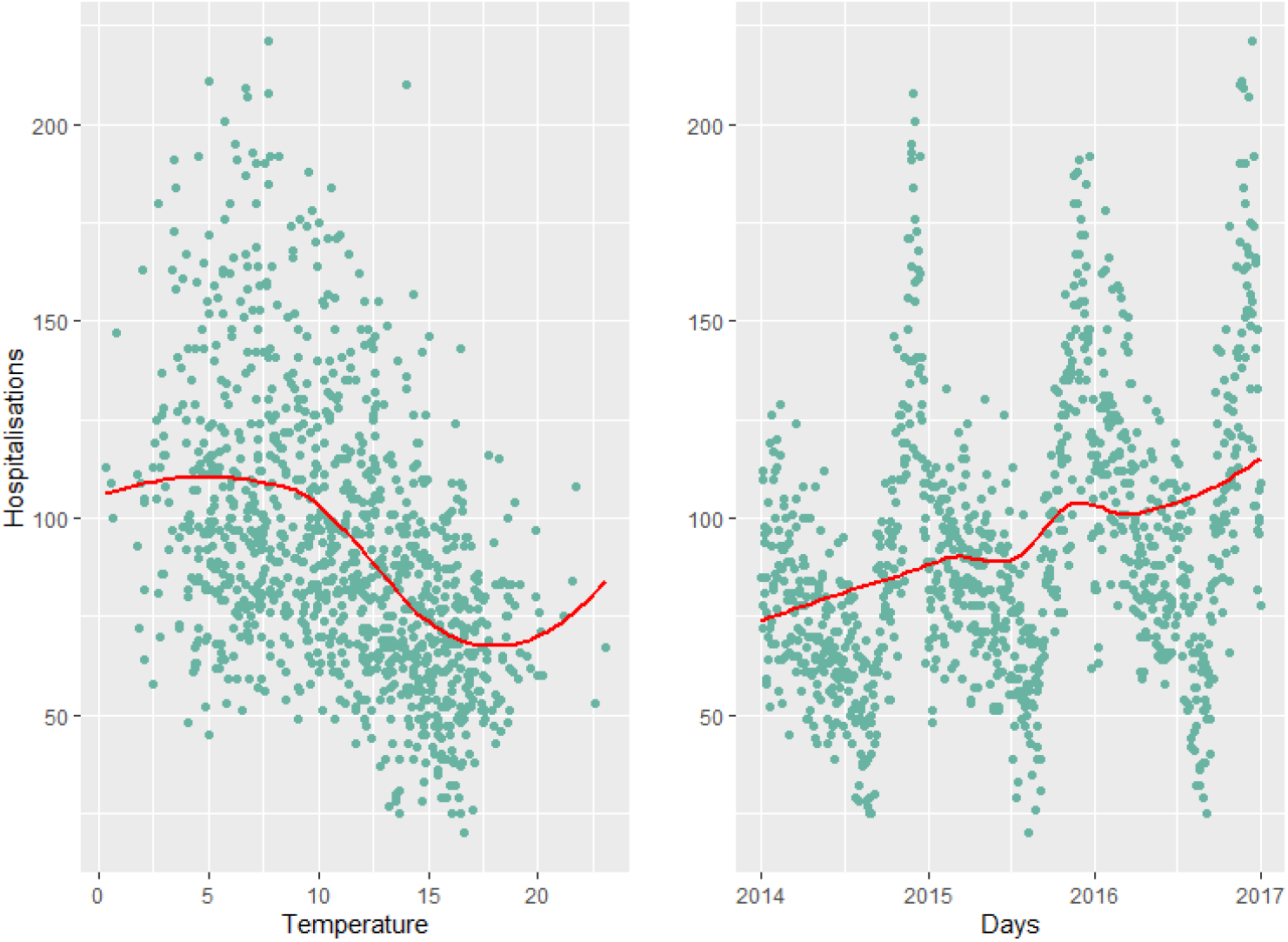
Relationship between mean daily temperature and paediatric hospital admissions (left); time series of paediatric hospital admissions (right). The red line represents the best fitting spline showing non-linearity between temperature (time) and hospital admissions.

### 3.1 Model selection

We ran the multivariate factor model multiple times, specifying a different number of factors (sources), ranging from 3 to 6, with the aim of finding the model most supported by the data. Figure 3 presents the plot of the Bayesian p-values and the *R*^2^ values for each model. The ones with 4 and 5 sources have the same value for R^2^ but the p-values are more in line with a uniform distribution for the model with 4 sources. To better discriminate between these two solutions, we also considered the plausibility of the profiles, i.e. their ability to cover the entire range of particle sizes, as well as the correlation with other pollutants and metereological variables. We did not find substantial differences in terms of correlation (Figure 5 here and Figure 1 in Supplementary Material); however, considering 5 factors seems to lead to a split of one source found in the 4 factor solution into two, resulting in a large overlap with another one. Due to this and the slightly better behaviour from a statistical perspective, we chose the solution with 4 sources as out final model.

**Figure 3.**
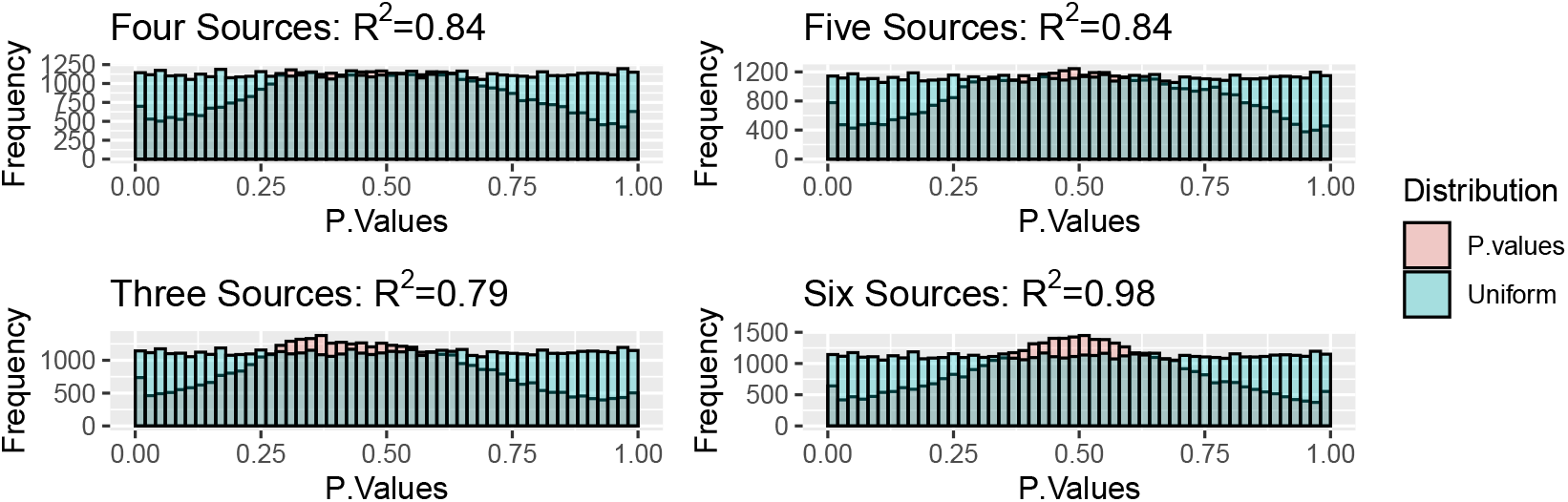
Model selection metrics

**Figure 4.**
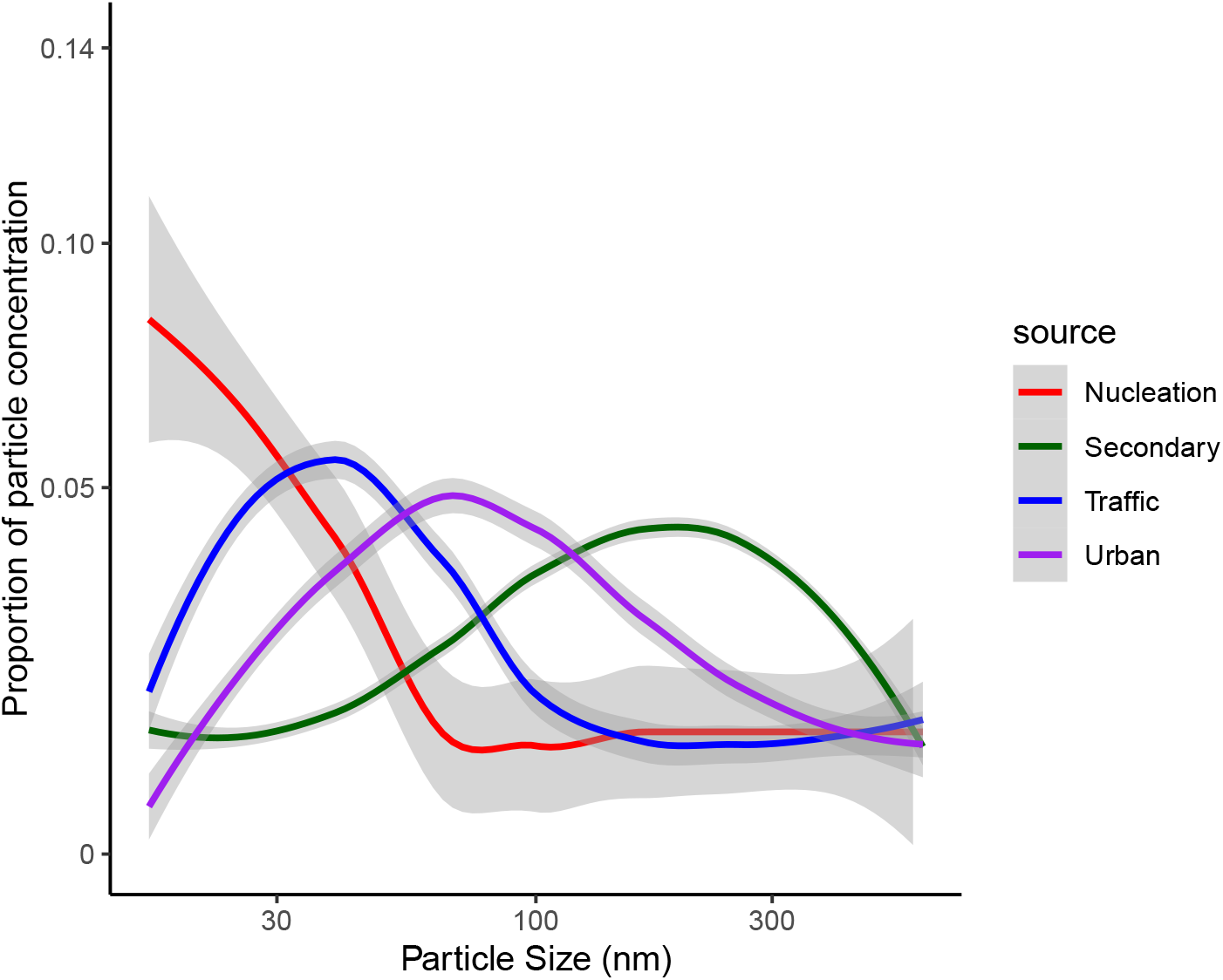
Results of Particle size distribution for the 4 sources identified by the model.

**Figure 5.**
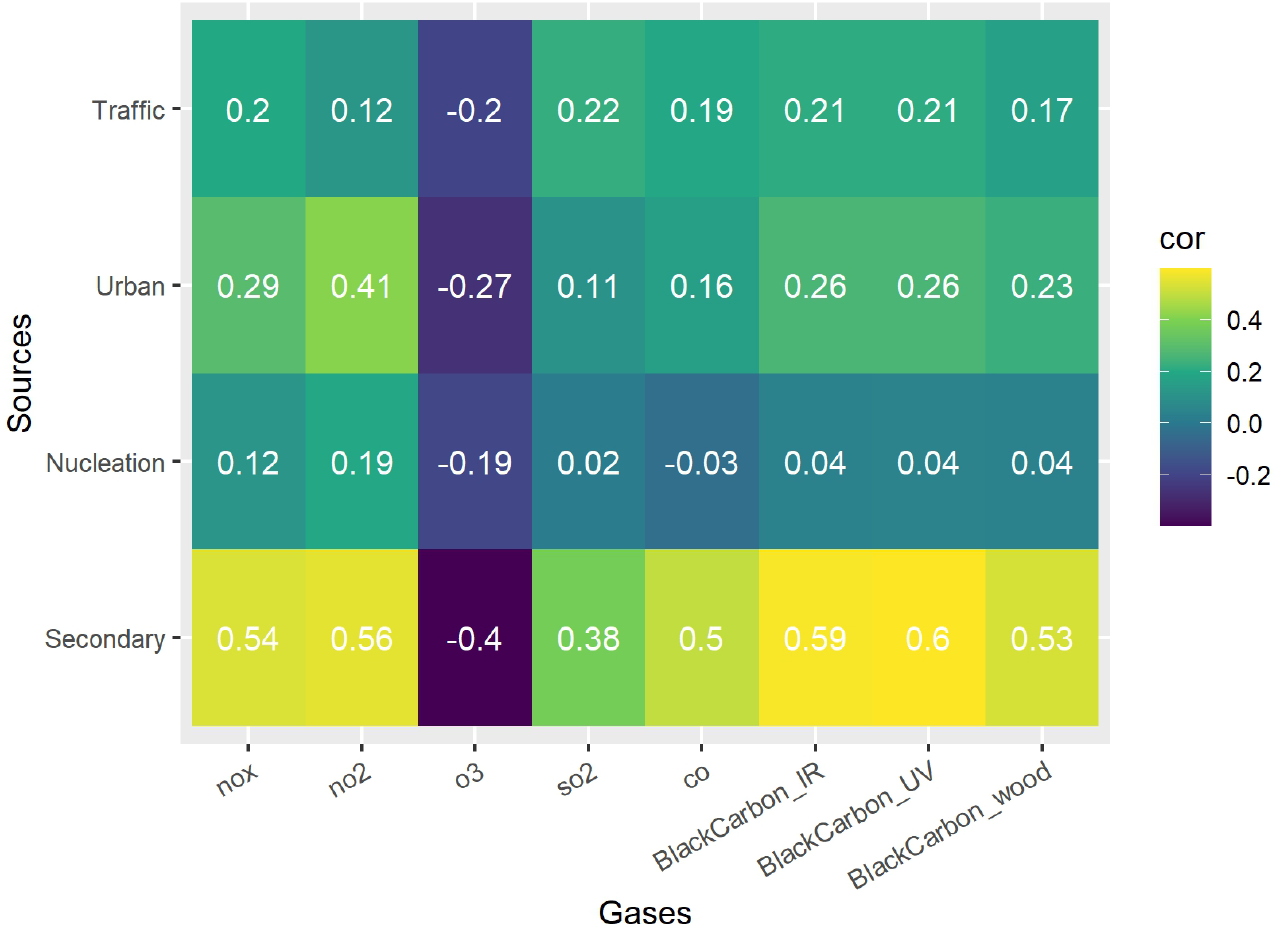
Correlation between the estimated sources and other air pollutants

### 3.2 Bayesian dynamic factor model results

The next step consists in interpreting the sources, giving them a label based on their profiles, their temporal patterns and their correlation with other gasses and metereological variables. We present source profiles in Figure 4, consisting of the proportion of particle concentration by bin sizes for each source.

Source 1 is characterized by a high concentration of particles in the smaller bins, ranging from 19.5 to 42.5 nm (mode equals to 19.5nm). We label this source as **nucleation**, referring to the process by which tiny particles form from gaseous precursors in the atmosphere. It could represent emissions from Heathrow Airport, particularly due to the prevalence of strong south-westerly winds (Figure 7). This source is more prevalent during summer and winter (Figure 6), but it does not show a strong correlation with any of the gases or meteorological variables (Figure 5). The estimated autoregressive parameter for Source 1, *ρ*_1_, was 0.957 (95% CI: 0.933–0.983).

**Figure 6.**
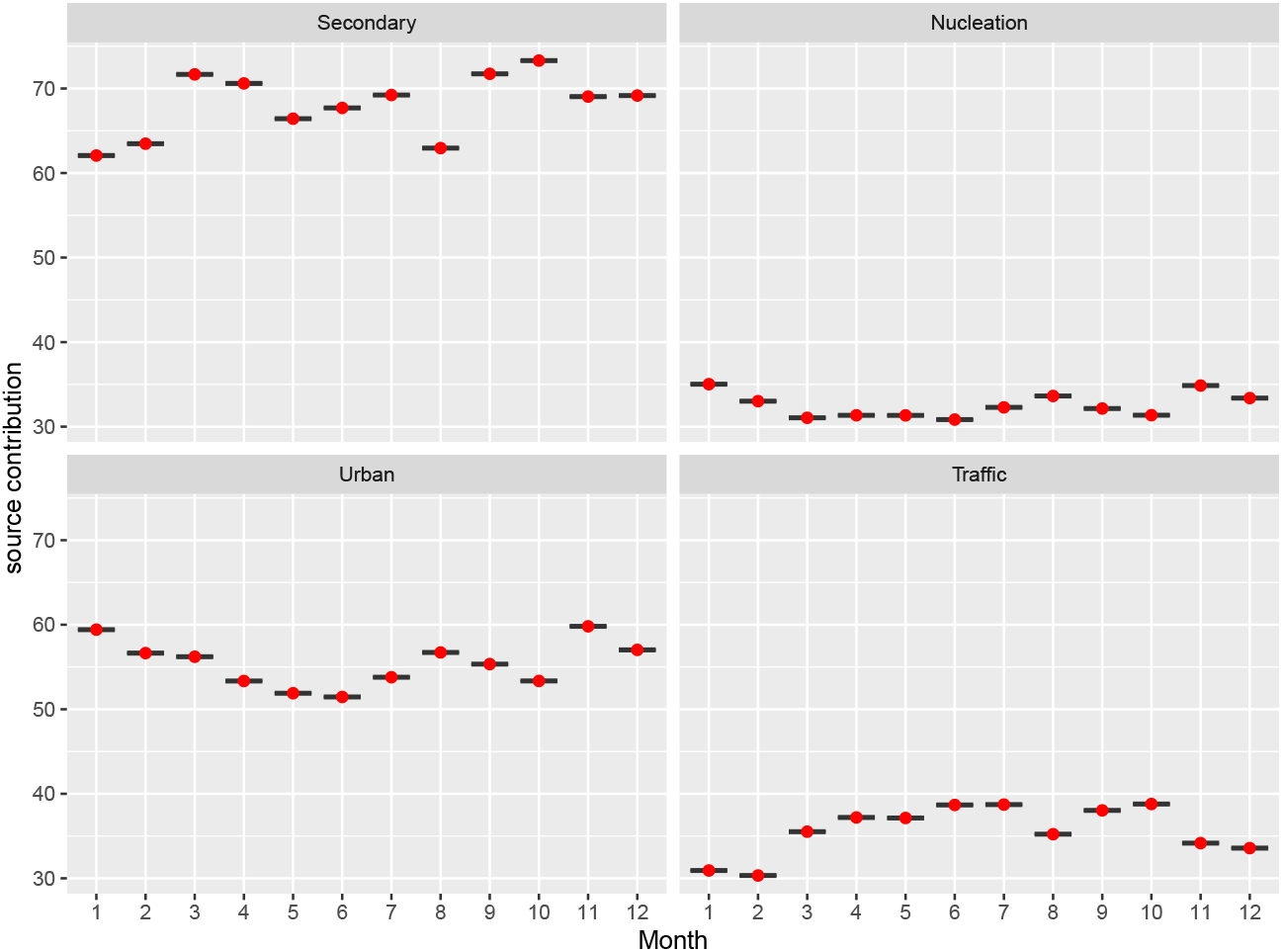
Estimated sources and average contributions per month

**Figure 7.**
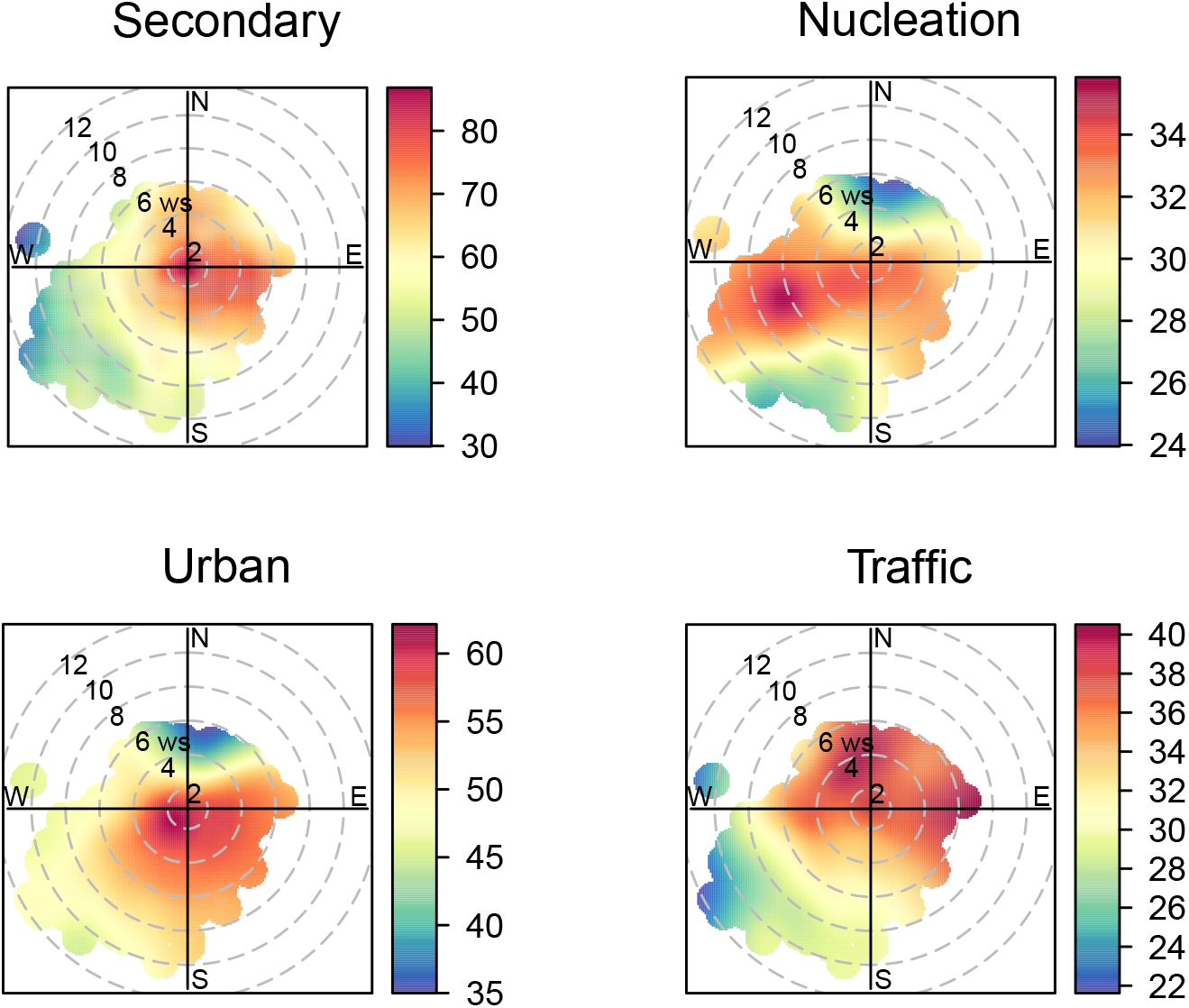
Polar plots showing how the estimated sources are affected by daily wind direction and wind velocity. The concentration scale of each source is reported in the legend.

Source 2 has the highest particle concentration of sized around 48-563nm (mode equals to 273.90 nm). This source shows strong seasonality, with peaks in March-April and then in September-October (—Figure 6 and has the strongest correlation with NO_*x*_, NO_2_ and CO, but also a negative correlation with visibility and O_3_. It is characterised by a weak wind coming from East (Figure 7). We label this **secondary**, as it could represent particles that are not directly emitted but are formed in the atmosphere through chemical reactions between primary pollutants and other atmospheric compounds, for instance from reactions between sulfur dioxide, nitrogen oxides, and ammonia. The estimated autoregressive parameter for source 2, *ρ*_2_, was of 0.979 (95%CI: 0.962 - 0.996). Source 3 has the highest particle concentration of bins sized around 20.55-93.05nm (mode equals to 45.30nm), with values close to 0 from 100nm and above. This source is slightly lower during winter and shows a moderate correlation with CO, NO_*x*_, SO_2_ and atmospheric pressure. Looking at the wind, we can see that it is charasterised by low speed and a North-East direction (Figure 7). We label this **traffic**, as is likely to represent the emissions coming from vehicles. The estimated autoregressive parameter for source 3, *ρ*_3_, was of 0.965 (95%CI: 0.942 - 0.988).

Source 4 has a peak in the particle concentration of sized between 31.65 and 254.90(mode equals to 75.05nm), but is characterised by a large variability. This source is lower in Spring and correlated moderately with NO_2_ (positively) and with O_3_ (negatively). It is characterised by Easterly wind (Figure 7) and we label this **urban**, as this behavior is consistent with emissions arising from urban activities besides transportation, for instance including heating and cooking, waste burning, energy production, constructions and demolitions and chemical uses in households and businesses. The estimated autoregressive parameter for source 4, *ρ*_4_, was of 0.974 (95%CI: 0.955 - 0.994).

### 3.3 Health model: phase 2

Figure 8 presents the point estimates and 95% credible intervals (95%CI) for the log relative risk of the four sources and lag 0 to 5. The green lines represent the results from the model without uncertainty (plugged-in), where we included the posterior mean of the daily sources contributions as exposure, while the orange lines visualise the results for the model with uncertainty propagated, obtained repeating the analysis 100 times, each time drawing and weighting the values of the four sources from their distributions obtained from the phase 1 model (weighted).

**Figure 8.**
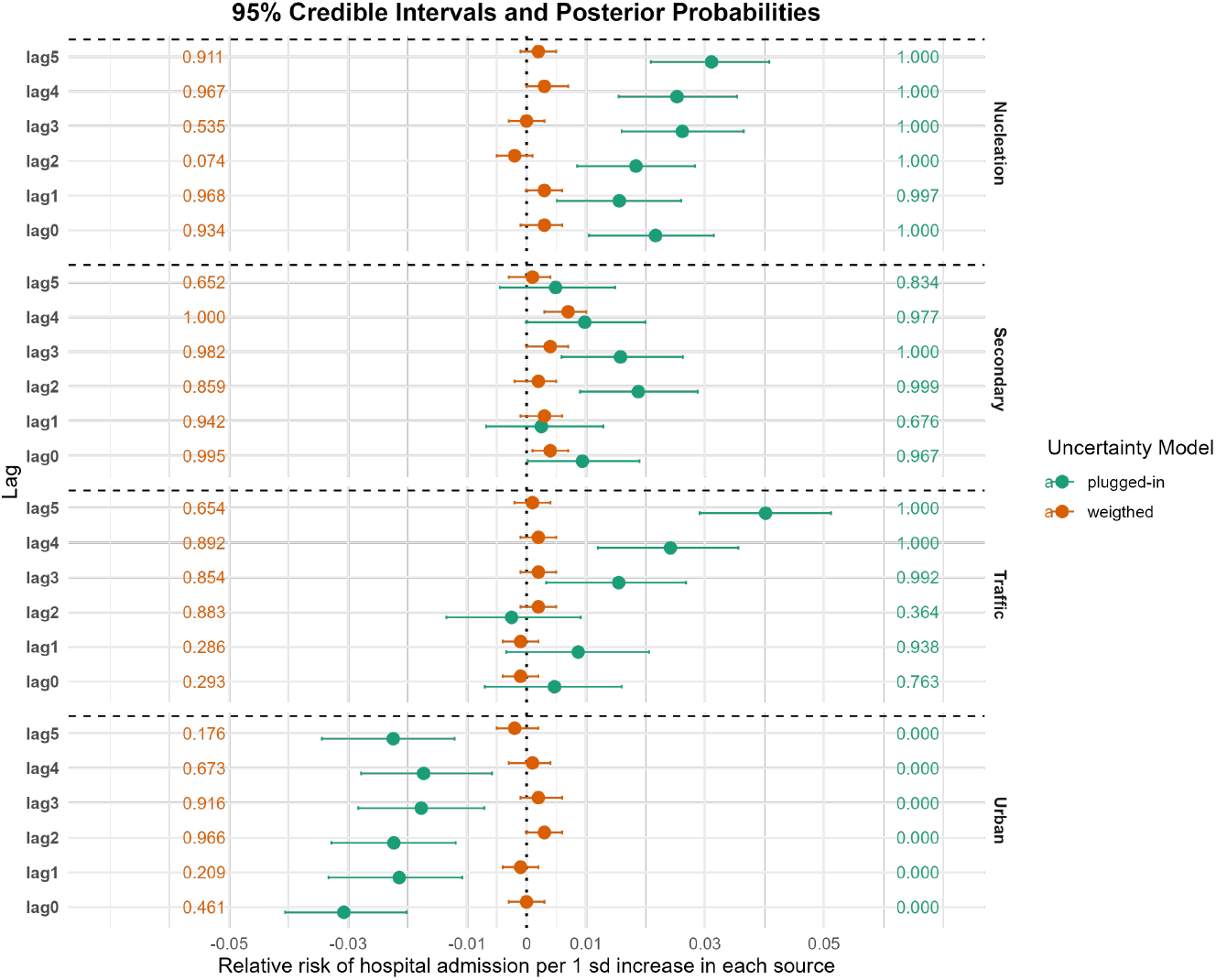
Results of the stage 2 model (plugged-in and weighted). The figure shows the log transformed relative risks and 95%CI for the lags 0 to 5 across the four air pollution sources for the model without uncertainty from stage 1 (plugged-in) (green lines) and with uncertainty propagated through a weighted combination of the posterior distributions (orange lines). Additionally, on the left and of the right end sides we present the posterior probability that the corresponding effect is above 0, indicating a positive association between the source and the risk of pediatric hospital admission in London.

Overall, there is also a shift towards 0 for most of the effects when uncertainty propagation is considered. Traffic and Secondary sources appear to have more consistent and positive effects, with high coefficients across lags, though these effects are lower when uncertainty is included. Nucleation and urban show the largest difference between the model without and with uncertainty. In the former, Nucleation has a positive association with risk of hospitalisations, which disappear completely when uncertainty is considered. Higher concentration of the Urban source is associated with lower risk of hospitalisation in the model without uncertainty across all the lags, while when uncertainty is propagated from the stage 1 model there is no evidence of an association anymore, except for lag 2 where the effect switch from negative to positive.

The comparison of the weighted model with the unweighted one, obtained combining the full posterior distributions of each element of the ***β*** using the *G* samples from the first stage model but without weighting them, is presented in Figure 9. Most of the point estimates are in agreement with the weigthed model ones, except for Nucleation, where across all lags the unweigthed model show negative estimates, while the weigthed ones are mostly on the positive side. Nevertheless, across all the sources and lags, the unweigthed approach is characterised by a large uncertainty, with likely values of ***β*** estimates ranging from −0.05 and 0.05.

**Figure 9.**
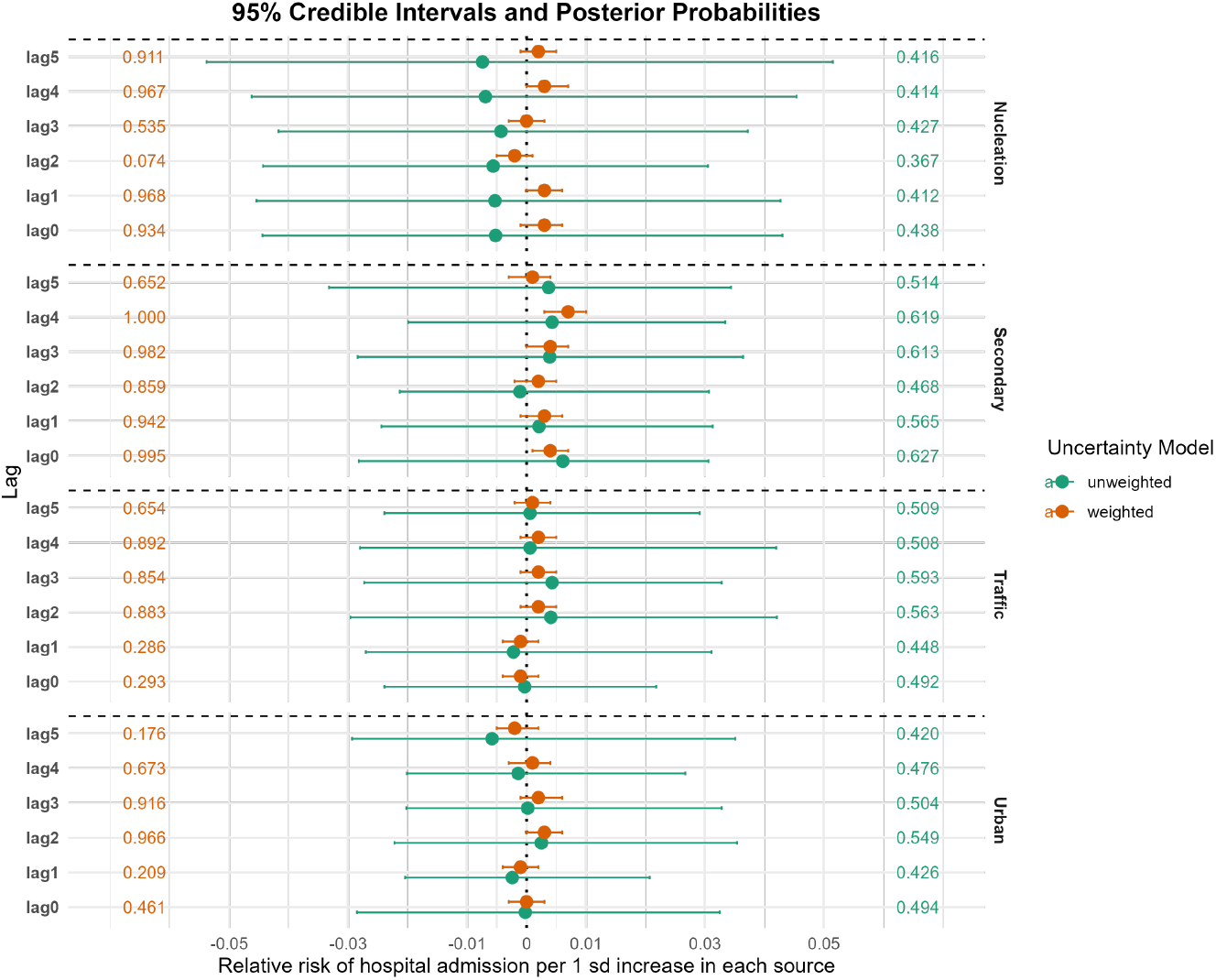
Results of the stage 2 model (unweighted and weighted). The figure shows the log transformed relative risks and 95%CI for the lags 0 to 5 across the four air pollution sources for the model where the uncertainty from stage 1 is incorporated through a simple pooling of the posterior distributions for each element in the ***β*** vector (unweighted - green lines) and through a weighted combination of the posterior distributions (weighted - orange lines). Additionally, on the left and of the right end sides we present the posterior probability that the corresponding effect is above 0, indicating a positive association between the source and the risk of pediatric hospital admission in London.

## 4 Discussion

This paper proposes a novel approach to SA and health risk assessment of air pollution using a two-stage Bayesian factor analysis. The methodology is applied to apportion PNSD and assess their health impacts. The first stage employs a Bayesian dynamic factor model with autoregressive components to estimate latent air pollution sources while enforcing nonnegativity constraints on source contributions and compositions. The second stage links these sources to respiratory hospital admissions in children through a Poisson regression model, with uncertainty from the first stage fully propagated into the health model.

COMEAP in its statement published in 2022 concluded that although the toxicological and epidemiological evidence shows that different constituents are likely to have different toxicological actions, there was no consistent indication of specific particle components that are more toxic than others; therefore they recommended research to address some outstanding issues, such as conducting epidemiological studies incorporating source apportionment data and designs of studies that can distinguish the effects of correlated exposures. This study aimed to address some of these by examining the intersection of air quality, health, and pollution sources, providing a science-driven pathway for achieving multiple SDGs while maintaining a strong focus on urban public health. In support of SDG 3, which focuses on good health and well-being, it highlights the impact of air pollution on child health. It also contributes to SDG 11, aimed at promoting sustainable cities and communities, by identifying key urban pollution sources and informing targeted mitigation strategies. The implementation of policies aiming to improve air quality (eg electric cars) has been affecting the particle composition in the atmosphere, and it would be important to understand the different health impacts of the various particle components. Lastly, the research emphasizes the disproportionate impact of air pollution on vulnerable populations, particularly children in urban areas, thereby contributing to SDG 10, which seeks to reduce inequalities.

Air pollution, especially PM, poses significant health risks, yet traditional epidemiological studies often overlook the heterogeneity of PM sources. By using PNSD data, which captures both ultrafine and fine particles, this study addresses a critical gap in source identification. A key advantage of the Bayesian approach is that it integrates uncertainty directly into the estimation of source profiles and contributions. In contrast, methods like PMF require the analyst to provide a priori estimates of measurement uncertainty to run the model. Also, while post hoc techniques such as bootstrapping can assess the stability of source profiles from PMF, in the Bayesian approach, we rely on an integrated treatment of uncertainty taking advantage of the full posterior distribution, which enhances the reliability of health impact estimates.

Through this analysis, four main pollution sources were identified: nucleation, traffic, urban activities, and secondary sources, each distinguished by unique particle size profiles and temporal patterns. These are in line with previous literature (Rivas et al., 2020; Garcia-Marlès et al., 2024). Among the apportioned sources, traffic and secondary factors exhibit the strongest and most consistent associations with respiratory health risks. Notably, the inclusion of uncertainty in the model highlights potential overestimates in traditional approaches that fail to incorporate uncertainty propagation.

These findings align with previous research on PNSD and health effects. Rivas et al. (2021) assessed the impact of ultrafine sources on daily mortality in Barcelona, Helsinki, London, and Zurich. In London, they found associations between nucleation, secondary sources, and traffic emissions with respiratory outcomes. However, in our study, when models were run without propagation of uncertainty, an unexpected inverse relationship (negative estimates) was found for urban sources, which disappeared when uncertainty was considered, emphasizing the importance of accounting for uncertainty in epidemiological models. Braniš et al. (2010) analyzed the association of particle number and PM_2.5_ concentrations with mortality and cardiorespiratory hospital admissions in Prague while Halonen et al. (2009) examined associations between different particle size modes and mortality/hospital admissions among 65+ in Helsinki, Finland. While in these studies no association was found between air pollution and daily mortality, Braniš et al. (2010) identified a link between concentration of submicron particles 15–487nm and respiratory admissions.

Our findings underscore the need for targeted mitigation strategies, particularly for vehicular emissions and secondary pollutants. The proposed methodological framework is adaptable to other regions and pollutants, providing a versatile tool for air quality management and health risk assessment.

Like most studies, this research has several limitations. One key consideration is the reliance on specific modeling assumptions. Although uncertainty has been accounted for, some significant associations may still be due to chance. The exposure model has been run on daily time series, while the typical approach in the source apportionment context would be to use hourly data, which are able to fully exploit the temporal variability in the air pollution concentration. However, our choice was dictated by the health data, which are only available at a daily resolution, making it not feasible to model hourly data. This could have an impact on the correlation between the sources and the pollutants or meteorological variables, resulting in values closer to zero. Another limitation pertains to exposure misclassification, as the data are derived from a single monitoring station, which may not fully capture the spatial heterogeneity of ultrafine particles. This limitation could affect the accuracy of health risk estimates. In addition, the temporal and spatial variability of air pollution further complicates the interpretation of health risk estimates, particularly in large urban environments where pollution levels fluctuate considerably.

To address these limitations, future studies should (i) develop an approach which allows to estimate sources using hourly data which can then be upscaled at the daily level in the epidemiological model; (ii) incorporate data from multiple monitoring stations to better account for spatial variability. Additionally, expanding the analysis to include additional health outcomes, such as asthma and cardiovascular diseases, could provide a more comprehensive understanding of the health effects of air pollution. Furthermore, investigating long-term exposure effects and interactions between different particle sources may yield deeper insights into their cumulative impacts on public health.

This paper advances air pollution research by integrating cutting-edge Bayesian techniques with real-world health data. The findings provide valuable insights for policymakers and establish a strong foundation for future interdisciplinary studies in environmental health.

## Acknowledgments

The work of the UK Small Area Health Statistics Unit is overseen by UK Health Security Agency (UK HSA). SAHSU is part of the MRC Centre for Environment and Health supported by the UK Medical Research Council, Grant number: MR/L01341X/1), and the National Institute for Health Research (NIHR) through its Health Protection Units (HPRUs) at Imperial College London in Environmental Exposures and Health and in Chemical and Radiation Threats and Hazards, and through Health Data Research UK (HDR UK) in the Social and Environmental Determinants of Health Driver Programme. This paper does not necessarily reflect the views of UKHSA, NIHR or the Department of Health and Social Care.

This work uses data provided by patients and collected by the NHS as part of their care and support. Hospital Episode Statistics data are copyright © 2025, re-used with the permission of NHS England. All rights reserved.

All authors acknowledge infrastructure support for the Department of Epidemiology and Biostatistics provided by the NIHR Imperial Biomedical Research Centre (BRC).

## Contributions

GB: methodology; analysis; writing (draft) MP: methodology; supervision; writing (draft); funding acquisition CM: writing (review); funding acquisition DG: data; writing (review); funding acquisition GF: data; writing (review); funding acquisition AT: data; writing (review); funding acquisition MB: methodology; supervision; writing (draft); funding acquisition

## Ethics

All data used in this study were taken from existing datasets and no original data from individuals or human samples were collected. Hospital admission data are already held by SAHSU, supplied by NHS digital; data use is covered by approval from national Research Ethics Services(reference 17/LO/0846) and by the Health Research Authority’s Confidentiality and Advisory Group (CAG) approval for section 251 support(reference 14/CAG/1039).

SAHSU holds approvals both from the London - South East Research Ethics Committee (22/LO/0256) and from the Health Research Authority - Confidentiality Advisory Group (20/CAG/0028).

## Patient consent for publication

Not required.

## Data availability

Due to confidentiality of the patient and ethical restrictions, the health data used in this study may be available only under specific conditions. However, the air pollution-related data sets analyzed during the current study are available from the corresponding author upon reasonable request.

## Footnotes

1 https://www.gov.uk/government/groups/committee-on-the-medical-effects-of-air-pollutants-comeap

## References

Baerenbold, O., M. Meis, I. Martínez-Hernández, C. Euán, W. S. Burr, A. Tremper, G. Fuller, M. Pirani, and M. Blangiardo (2023). A dependent bayesian dirichlet process model for source apportionment of particle number size distribution. Environmetrics 34 (1), e2763.

Beddows, D. and R. M. Harrison (2019). Receptor modelling of both particle composition and size distribution from a background site in london, uk–a two-step approach. Atmospheric Chemistry and Physics 19 (7), 4863–4876.

Beddows, D., R. M. Harrison, D. Green, and G. Fuller (2015). Receptor modelling of both particle composition and size distribution from a background site in london, uk. Atmospheric Chemistry and Physics 15 (17), 10107–10125.

Bell, M. L., K. Ebisu, B. P. Leaderer, J. F. Gent, H. J. Lee, P. Koutrakis, Y. Wang, F. Dominici, and R. D. Peng (2014). Associations of pm2. 5 constituents and sources with hospital admissions: analysis of four counties in connecticut and massachusetts (usa) for persons 65 years of age. Environmental health perspectives 122 (2), 138–144.

Blangiardo, M., F. Finazzi, and M. Cameletti (2016). Two-stage bayesian model to evaluate the effect of air pollution on chronic respiratory diseases using drug prescriptions. Spatial and spatio-temporal epidemiology 18, 1–12.

Blangiardo, M., M. Pirani, L. Kanapka, A. Hansell, and G. Fuller (2019). A hierarchical modelling approach to assess multi pollutant effects in time-series studies. PLoS One 14 (3), e0212565.

Boogaard, H., A. Patton, R. Atkinson, J. Brook, H. Chang, D. Crouse, J. Fussell, G. Hoek, B. Hoffmann, R. Kappeler, M. Kutlar Joss, M. Ondras, S. Sagiv, E. Samoli, R. Shaikh, A. Smargiassi, A. Szpiro, E. Van Vliet, D. Vienneau, J. Weuve, F. Lurmann, and F. Forastiere (2022). Long-term exposure to traffic-related air pollution and selected health outcomes: A systematic review and meta-analysis. Environment International 164, 107262.

Braniš, M., J. Vyškovská, M. Malý, and J. Hovorka (2010). Association of size-resolved number concentrations of particulate matter with cardiovascular and respiratory hospital admissions and mortality in prague, czech republic. Inhalation toxicology 22 (Sup2), 21–28.

Carslaw, D. (2023). worldmet: Import surface meteorological data from NOAA integrated surface database (ISD). R package version 0.9.8.

Carslaw, D. C. and K. Ropkins (2012). openair — an R package for air quality data analysis. Environmental Modelling & Software 27–28 (0), 52–61.

COMEAP (2022). Particulate air pollution: health effects of exposure.

Crainiceanu, C. M., D. Ruppert, and M. P. Wand (2005). Bayesian analysis for penalized spline regression using winbugs. Journal of statistical software 14, 1–24.

Evangelopoulos, D., K. Katsouyanni, H. Walton, and M. Williams (2019). Personalising the health impacts of air pollution: Interim statistics summary for a selection of statements. King’s College London.

Finley, A. O. (2013). Using jags in r with the rjags package.

Font, A., K. Ciupek, D. Butterfield, and G. Fuller (2022). Long-term trends in particulate matter from wood burning in the united kingdom: Dependence on weather and social factors. Environmental Pollution 314, 120105.

Fuller, G. (2019). The invisible killer: The rising global threat of air pollution- and how we can fight back. Melville House.

Fuller, R., P. J. Landrigan, K. Balakrishnan, G. Bathan, S. Bose-O’Reilly, M. Brauer, J. Caravanos, T. Chiles, A. Cohen, L. Corra, et al. (2022). Pollution and health: a progress update. The Lancet Planetary Health 6 (6), e535–e547.

Garcia-Marlès, M., R. Lara, C. Reche, N. Pérez, A. Tobías, M. Savadkoohi, D. Beddows, I. Salma, M. Vörösmarty, T. Weidinger, C. Hueglin, N. Mihalopoulos, G. Grivas, P. Kalkavouras, J. Ondracek, N. Zikova, J. V. Niemi, H. E. Manninen, D. C. Green, A. H. Tremper, M. Norman, S. Vratolis, E. Diapouli, K. Eleftheriadis, F.J. Gómez-Moreno, E. Alonso-Blanco, A. Wiedensohler, K. Weinhold, M. Merkel, S. Bastian, B. Hoffmann, H. Altug, J.-E. Petit, P. Acharja, O. Favez, S. M. D. Santos, J.-P. Putaud, A. Dinoi, D. Contini, A. Casans, J. A. Casquero-Vera, S. Crumeyrolle, E. Bourrianne, M. V. Poppel,F. E. Dreesen, S. Harni, H. Timonen, J. Lampilahti, T. Petäjä, M. Pandolfi, P. K. Hopke, R. M. Harrison, A. Alastuey, and X. Querol (2024). Source apportionment of ultrafine particles in urban europe. Environment International 194, 109149.

Gelman, A., J. B. Carlin, H. S. Stern, D. B. Dunson, A. Vehtari, and D. B. Rubin (2013). Bayesian data analysis. Chapman and Hall/CRC.

Gu, J., M. Pitz, J. Schnelle-Kreis, J. Diemer, A. Reller, R. Zimmermann, J. Soentgen, M. Stoelzel, H.-E. Wichmann, A. Peters, et al. (2011). Source apportionment of ambient particles: comparison of positive matrix factorization analysis applied to particle size distribution and chemical composition data. Atmospheric Environment 45 (10), 1849–1857.

Hackstadt, A. J. and R. D. Peng (2014). A bayesian multivariate receptor model for estimating source contributions to particulate matter pollution using national databases. Environmetrics 25 (7), 513–527.

Halonen, J. I., T. Lanki, T. Yli-Tuomi, P. Tiittanen, M. Kulmala, and J. Pekkanen (2009). Particulate air pollution and acute cardiorespiratory hospital admissions and mortality among the elderly. Epidemiology 20 (1), 143–153.

Hopke, P. K. (2016). Review of receptor modeling methods for source apportionment. Journal of the Air & Waste Management Association 66 (3), 237–259.

Lee, D., S. Mukhopadhyay, A. Rushworth, and S. K. Sahu (2017). A rigorous statistical framework for spatio-temporal pollution prediction and estimation of its long-term impact on health. Biostatistics 18 (2), 370–385.

Martínez-Hernández, I., C. Euán, W. S. Burr, M. Meis, M. Blangiardo, and M. Pirani (2025). Modelling particle number size distribution: a continuous approach. Journal of the Royal Statistical Society Series C: Applied Statistics 74 (1), 229–248.

Molitor, J., E. Coker, M. Jerrett, B. Ritz, A. Li, et al. (2016). Part 3. modeling of multipollutant profiles and spatially varying health effects with applications to indicators of adverse birth outcomes. Research report (Health Effects Institute) (183 Pt 3), 3–47.

Park, E. S., P. Guttorp, and R. C. Henry (2001). Multivariate receptor modeling for temporally correlated data by using MCMC. Journal of the American Statistical Association 96 (456), 1171–1183.

Park, E. S., P. K. Hopke, M.-S. Oh, E. Symanski, D. Han, and C. H. Spiegelman (2014). Assessment of source-specific health effects associated with an unknown number of major sources of multiple air pollutants: a unified bayesian approach. Biostatistics 15 (3), 484–497.

Park, E. S., E.-K. Lee, and M.-S. Oh (2021). Bayesian multivariate receptor modeling software: Bnfa and bayesmrm. Chemometrics and Intelligent Laboratory Systems 211, 104280.

Park, E. S. and M.-S. Oh (2015). Robust bayesian multivariate receptor modeling. Chemometrics and Intelligent Laboratory Systems 149, 215–226.

Park, E. S., E. Symanski, D. Han, and C. Spiegelman (2015). Part 2. development of enhanced statistical methods for assessing health effects associated with an unknown number of major sources of multiple air pollutants. Research report (Health Effects Institute) (183 Pt 1-2), 51–113.

Pirani, M., N. Best, M. Blangiardo, S. Liverani, R. W. Atkinson, and G. W. Fuller (2015). Analysing the health effects of simultaneous exposure to physical and chemical properties of airborne particles. Environment international 79, 56–64.

Pirani, M., A. Panton, D. A. Purdie, and S. K. Sahu (2016). Modelling macronutrient dynamics in the hampshire avon river: A bayesian approach to estimate seasonal variability and total flux. Science of the Total Environment 572, 1449–1460.

Rivas, I., D. C. Beddows, F. Amato, D. C. Green, L. Järvi, C. Hueglin, C. Reche, H. Timonen, G. W. Fuller, J. V. Niemi, et al. (2020). Source apportionment of particle number size distribution in urban background and traffic stations in four european cities. Environment international 135, 105345.

Rivas, I., L. Vicens, X. Basagaña, A. Tobías, K. Katsouyanni, H. Walton, C. Hüglin, A. Alastuey, M. Kulmala, R. M. Harrison, et al. (2021). Associations between sources of particle number and mortality in four european cities. Environment International 155, 106662.

Ruppert, D., M. Wand, and R. Carroll (2003). Semiparametric Regression. Cambridge University Press.

Samoli, E., R. W. Atkinson, A. Analitis, G. W. Fuller, D. Beddows, D. C. Green, I. S. Mudway, R. M. Harrison, H. R. Anderson, and F. J. Kelly (2016). Differential health effects of short-term exposure to source-specific particles in london, uk. Environment international 97, 246–253.

Szpiro, A. A. and C. J. Paciorek (2013). Measurement error in two-stage analyses, with application to air pollution epidemiology. Environmetrics 24 (8), 501–517.

Tang, J.-H., S.-C. C. Lung, and J.-S. Hwang (2020). Source apportionment of pm2. 5 concentrations with a bayesian hierarchical model on latent source profiles. Atmospheric Pollution Research 11 (10), 1715–1727.

Tremper, A. H., C. Jephcote, J. Gulliver, L. Hibbs, D. C. Green, A. Font, M. Priestman, A. L. Hansell, and G. W. Fuller (2022). Sources of particle number concentration and noise near london gatwick airport. Environment International 161, 107092.

Viana, M., T. Kuhlbusch, X. Querol, A. Alastuey, R. Harrison, P. Hopke, W. Winiwarter, M. Vallius, S. Szidat, A. Prevot, C. Hueglin, H. Bloemen, P. Wahlin, R. Vecchi, A. Miranda, A. Kasper-Giebl, W. Maenhaut, and R. Hitzenberger (2008). Source apportionment of particulate matter in Europe: a review of methods and results. Journal of Aerosol Science 39, 827–849.

Villejo, S. J., J. B. Illian, and B. Swallow (2023). Data fusion in a two-stage spatio-temporal model using the inla-spde approach. Spatial Statistics 54, 100744.

WHO (2021). WHO Global Air Quality Guidelines: Particulate Matter (PM2.5 and PM10), Ozone, Nitrogen Dioxide, Sulfur Dioxide and Carbon Monoxide. Geneva, Switzerland: World Health Organization.

WHO (2025). Air pollution and sustainable development goals (sdgs).

Wiedensohler, A., W. Birmili, A. Nowak, A. Sonntag, K. Weinhold, M. Merkel, B. Wehner, T. Tuch, S. Pfeifer, M. Fiebig, et al. (2012). Mobility particle size spectrometers: Harmonization of technical standards and data structure to facilitate high quality long-term observations of atmospheric particle number size distributions. Atmospheric Measurement Techniques 5 (3), 657–685.

Yu, L., W. Liu, X. Wang, Z. Ye, Q. Tan, W. Qiu, X. Nie, M. Li, B. Wang, and W. Chen (2022). A review of practical statistical methods used in epidemiological studies to estimate the health effects of multi-pollutant mixture. Environmental Pollution 306, 119356.

